# In Silico Modeling of Transcatheter Heart Valve Oversizing and Ellipticity, Part I: Establishing Credibility of an Advanced Model

**DOI:** 10.1101/2025.09.25.25336634

**Authors:** Sam Boxwell, Dylan Armfield, Rachel M.E. Cahalane, William Hickey, Scott Cook, Patricia Kelly, Philip Cardiff, Laoise McNamara

**Author notes:** **Corresponding Author:** Laoise McNamara.

## Abstract

**Background and Objectives:** Transcatheter aortic valve implantation (TAVI) is the most common modality of treatment for aortic stenosis. However, transcatheter heart valves (THVs) can be prone to early failure and an increase in thrombogenic events, yet the risk factors associated with these failure modes remain poorly understood. Computational modeling may be used to predict biomechanical indices associated with degeneration and thrombogenicity, however existing models do not fully account for complex stent and leaflet material behavior, and establishing model credibility according to ASME VV-40 is required.

**Methods:** In this study, we developed an advanced structural and hemodynamic *in silico* framework to predict the *in vitro* performance of a supra-annular, self-expanding THV across a range of clinically-relevant expansion and ellipticity indices. The THV was modelled by incorporating a novel 3-fibre material model for pericardium tissue leaflets and a super-elastic nitinol stent.

**Results:** Calculation verification was conducted and, on this basis, we provide recommendations on mesh density, element integration and target time increment. Following verification, we validated our models with radial force, structural camera and hemodynamic particle image velocimetry testing across multiple THV deployment configurations. In the ‘min-oversizing, circular’ case, we predicted a similar geometric orifice area (4.35 vs 4.02cm^2^), pinwheeling index (2.6% vs 2.7%), stent deflection (1.95 vs 1.76mm) and flow velocity (1.33 vs 1.27m/s) to *in vitro* data.

**Conclusion:** Thus, we validated a novel structural and hemodynamic *in silico* framework for studying THVs, which will be applied to understand deployment factors contributing to structural degeneration and thrombogenicity. This framework also holds potential for guiding next-generation THV design and predictive procedural modeling.

## 1 Introduction

Aortic stenosis, the most prevalent valvular heart disease in developed countries, is marked by progressive fibro-calcific remodeling of aortic valve leaflets, which reduces leaflet motion and restricts blood flow, eventually resulting in hypertrophy and heart failure [1]. Until recently, treatment of this condition involved surgical aortic valve replacement (SAVR), whereby the diseased native valve was removed and replaced with a bioprosthetic device through open heart surgery. However, transcatheter aortic valve implantation (TAVI) has been developed as a minimally invasive alternative to SAVR, involving the crimping of a bioprosthetic device onto a catheter, introduced percutaneously and deployed over the diseased native valve. TAVI has now become the most common treatment modality for patients with aortic stenosis, due to proven efficacy in high, intermediate and low-risk patient cohorts [2, 3]. However, complications can arise during and after the procedure including paravalvular leakage, conduction interference and structural valve degeneration [4-6]. These complications are intrinsically linked to interactions between the native anatomy, the transcatheter heart valve (THV) and adjacent blood flow, but further understanding of these interactions is necessary for optimizing device design and improving clinical outcomes.

Due to the challenges of studying device design and deployment in bench top-models, pre-clinical animal models and clinical trials, computational modeling has been used for biomechanical assessment of THV performance across the device lifecycle; from concept development, pre-operative planning and post-operative assessment [7-9]. Previous studies have utilized computational solid mechanics [10-13], computational fluid dynamics (CFD) [14-16] or more recently, fluid-structure interaction (FSI) [7, 17, 18] approaches, and often simulate THV deployment within idealized aortic roots [7, 11, 19] or patient-specific approaches involving reconstruction from pre-TAVI computed tomography [11-13]. Such approaches offer valuable insights into THV performance *in vivo* and may be used to predict the onset of paravalvular leakage [14], conduction interference [12] and structural valve degeneration [7, 15, 16]. Despite this, there are still notable discrepancies between modeling approaches of THVs with respect to geometry, material properties and element formulation. Although it is well known that glutaraldehyde-fixed pericardium is both hyperelastic and anisotropic, with numerous studies exploring the influence of both collagen fiber orientation and architecture on the influence of valve dynamics [20-22], bioprosthetic leaflets of THVs have been modelled as linear-elastic [13, 17, 23], isotropic hyperelastic [11, 24, 25] or anisotropic hyperelastic materials [7, 10, 18, 21]. Furthermore, while many computational studies neglect leaflet-frame interaction by assuming stent rigidity post-THV deployment [9, 11, 19], our previous work showed that a new-generation self-expanding THV exhibited notable radial stent deflections during the cardiac cycle due to pulsatile loading [10]. These deflections have also been reported across other new-generation devices [26], but are rarely accounted for in computational modeling studies of THV function. Thus, there is a need to further advance computational modeling approaches to more accurately represent device components, so that such approaches may be used for next-generation device design and development.

Establishing the reliability of computational models is a key prerequisite before such models may be adopted in clinical practice. The American Society of Mechanical Engineers (ASME) developed a risk-informed credibility assessment framework known as the ‘Verification and Validation in Computational Modeling of Medical Devices’ (VV 40-2018) [27]. This framework begins by defining the question of interest and the context of use (COU) to outline the scope and the role the computational model will play in the decision-making process. A credibility assessment is conducted, whereby the level of verification and validation required is proportional to the model risk, which in turn, is informed by the model influence and the consequences of an incorrect model prediction. In this context, verification is the process of ensuring correct mathematical implementation, while validation determines the model accuracy for representing real world application. This risk-informed framework has been applied for verification and validation of computational models of thrombectomy [28], thoracic endovascular aortic repair [29] and femoral artery stents [30]. During a credibility assessment while developing finite element (FE) models of a femoral artery stent, Bernini et al. showed that model outputs relating to crimping, deployment and fatigue prediction are highly sensitive to mesh discretization, element formulation, and target time increment [30]. Similarly, verification of mesh discretization and element formulation has been reported for heart valve dynamics, albeit excluding the THV stent frame [31]. Nonetheless, these studies highlight the need for careful sensitivity analysis of each of these numerical parameters to thereby ensure model reliability. In a more recent study, Grossi et al. showed successful development and validation of patient-specific FE models of various self-expanding THVs [13]. This study focused on calibration and validation of nitinol material models, which will directly impact crimping, deployment and conformance of the device with the native anatomy. Despite introducing a novel workflow for validating *in vivo* deployment of self-expanding THVs, this study did not account for cardiac function. Thus, there is a distinct need to establish model reliability of TAVI simulations across the entire device lifecycle, including crimping, deployment, cardiac cycle device dynamics and hemodynamics. These stages influence how the device will interact with the native environment *in vivo*, directly impacting patient-outcomes.

The present work is the first part of a comprehensive two-part study. The objectives of this first part are to (1) develop an advanced computational model of a THV to more accurately represent device components, and (2) conduct a credibility analysis of a computational solid mechanics and CFD model of a commercial THV based on the framework outlined in ASME-VV-40. Based on this, we conduct a range of verification and validation activities for (1) crimping and deployment, (2) cardiac cycle device dynamics and (3) cardiac cycle device hemodynamics. Thus, we establish, for the first-time, a novel workflow, encompassing all stages of the device implantation and deployment lifecycle, for verification and validation of TAVI simulations. In part two of this study, we use this validated approach to address our question of interest which aims to investigate the impact of clinically-relevant deployment configurations on the impact of device dynamics and degeneration [32].

## 2 Methods

### 2.1 Numerical model of THV

FE models of the ACURATE Prime XL THV (Boston Scientific, Galway, Ireland) were developed using Abaqus/Explicit (v2023, Simulia, Dassault Systémes, Providence, RI, USA). The device model included the porcine pericardium leaflets, an inner sealing skirt, the outer skirt and the nitinol stent, with relevant device geometries obtained from Boston Scientific. Similar to our previous approach [10], we simulated fabrication to initially attach a planar half-leaflet assembly (consisting of a half-leaflet and inner sealing skirt) to the stent and replicate the suturing process *in silico*, see Figure 1(a). This involved: (1) displacement of the leaflet commissures and edge of the inner sealing skirt to their respective suturing position, (2) application of a pressure-based loading condition to the surface of the half-leaflet assembly to push this component radially outwards until it contacted a cylindrical surface, and (3) application of another, small pressure-based loading condition to the half-leaflet assembly to further push this component radially outwards until it contacted the stent of the ACURATE Prime XL, which as part of this initial fabrication simulation was assumed to be a rigid structure. The resulting half-leaflet assembly was then exported as an orphan mesh and repeated in a circular pattern to form the fabricated tri-leaflet geometry. The stent and fabricated leaflets and inner skirt were fixed together with (1) node-to-surface tie constraints between the commissures and the stent, (2) surface-to-surface tie constraints between the aortic surface of the leaflets and the stent and (3) surface-to-surface tie constraints between the aortic surface of the inner skirt and the stent, where specific position tolerances were chosen to ensure that attachment occurred at suturing points. The geometry of the outer skirt was generated along nodal coordinates along the THV stent using Inventor (Autodesk, Inc., San Rafael, CA) and attached using a node-to-surface tie constraint. The THV stent was discretized using three-dimensional linear solid elements. As per ASME VV-40, we conducted verification analysis on the element formulation for the stent, evaluating the impact of full-, reduced-integration and kinematic split options (Section 2.3). To capture transmural behavior [33], the leaflet, inner sealing skirt and outer skirt were discretized with solid elements – specifically, hexahedral incompatible mode (C3D8I) elements with second order accuracy [11], with a global element size of 0.25 mm. Whilst further mesh refinement of the leaflet was required to crimp the device, an additional mesh sensitivity analysis also confirmed the need for three elements through the thickness of the leaflet for accurate stress analysis (Supplementary Section 1). The complete numerical model of the ACURATE Prime XL THV is shown in Figure 1(b).

**Figure 1:**
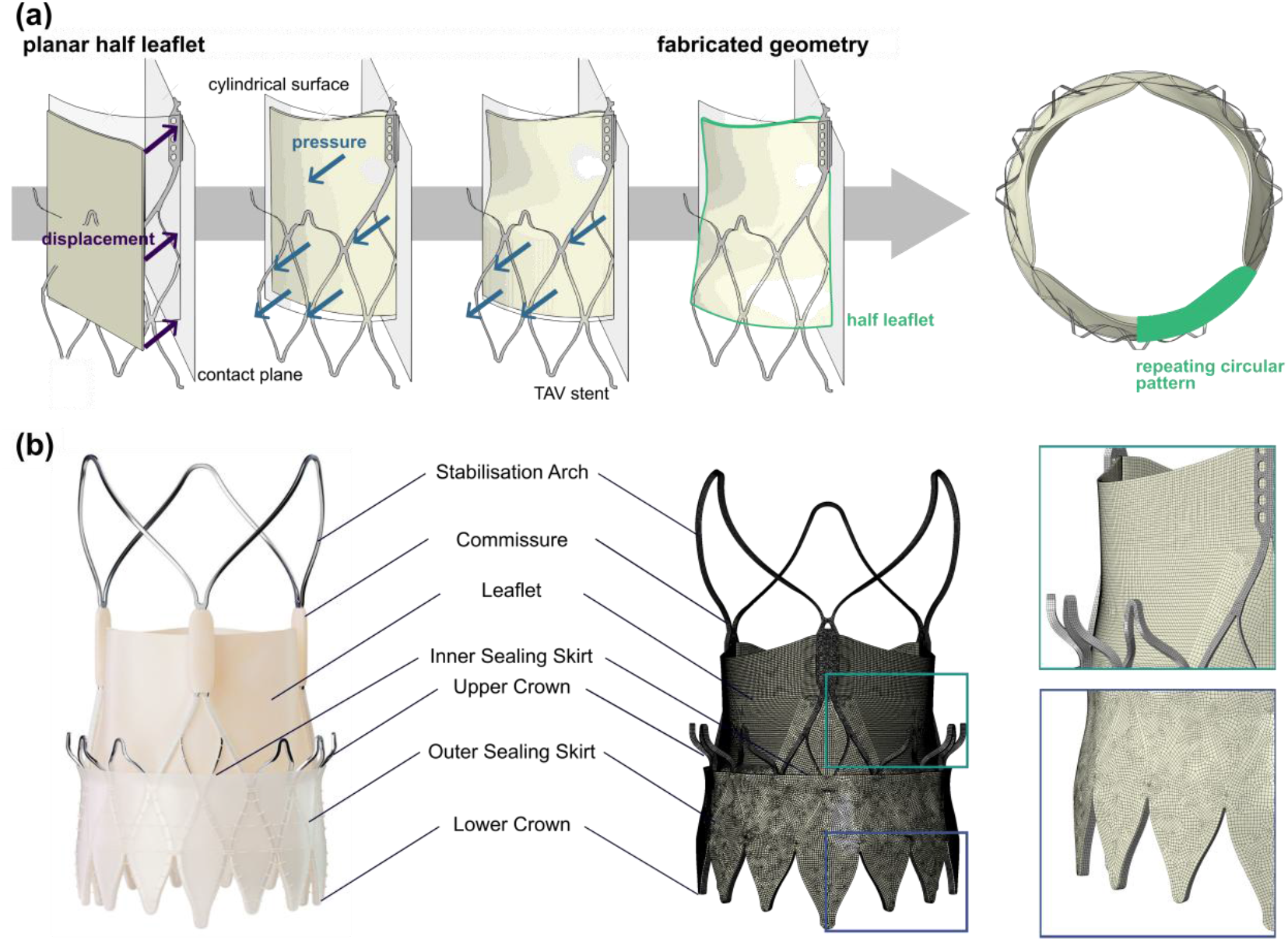
Finite element model of transcatheter heart valve (THV). (a) Simulation of THV fabrication to represent the attachment of the leaflet assembly to the stent. (b) Numerical model of ACURATE Prime XL showing mesh detail.

#### 2.1.1 Material modeling

##### 2.1.1.1 Glutaraldehyde-fixed porcine pericardium

In this study, we implemented a novel constitutive material model to describe the behavior of glutaraldehyde-fixed porcine pericardium, deemed the non-linear matrix, 3-fibre (NLM-3F) model. This model is extended from our previous approach [10], and describes the strain energy potential (*Ψ*), in terms of three components, a volumetric component (*Ψ*_*VOL*_), an isotropic isochoric component (*Ψ*_*ISO*_), representing the tissue matrix, and an anisotropic component (*Ψ*_*ANISO*_), representing the fibers within the porcine pericardium, which can be expanded as follows:

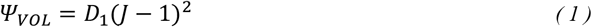

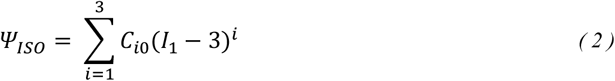

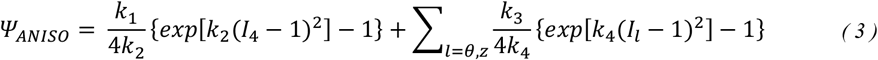

where *J* is the Jacobian determinant of the deformation gradient (***F***), *I*_1_ is the first invariant of the right Cauchy–Green deformation tensor and *I*_4_ and *I*_*l*_ are the invariants aligned in the direction of the collagen fibres. The isotropic component, which will control the out-of-plane response of the material, is modelled using the Yeoh formulation, as we postulate that the out-of-plane response of glutaraldehyde-fixed porcine pericardium will also exhibit strain-stiffening. To compute the NLM-3F model parameters, a least-squares fitting approach, with respect to the second Piola–Kirchhoff stress, was employed and fit to multi-protocol stress-strain curves of glutaraldehyde-fixed porcine pericardium obtained from Sulejmani et al. [34]. The fitting tool for this material model is openly available on <u>Github</u>. The resulting NLM-3F model approximations are presented in Figure 2(a), with corresponding parameters shown in Table 1. This was implemented to model glutaraldehyde-fixed porcine pericardium using an invariant-based strain energy potential function (*vuanisohyper_inv*), wherein fiber directions were defined as element local directions. Although the skirt was also composed of pericardium, we used the incompressible, hyperelastic, isotropic Marlow model for glutaraldehyde-fixed porcine pericardium of the outer skirt, which was fit to equibiaxial test data [34].

**Table 1:** Non-linear, 3-fibre (NLM-3F) material model parameters for glutaraldehyde-fixed porcine pericardium.

| $C_{10}$ (MPa) | $C_{20}$ (MPa) | $C_{30}$ (MPa) | $k_1$ (MPa) | $k_2$ | $k_3$ (MPa) | $k_4$ |
| --- | --- | --- | --- | --- | --- | --- |
| 0.05 | 4.13 | 4.86 | 1.00 | 65.97 | 1.32 | 58.64 |

**Figure 2:**
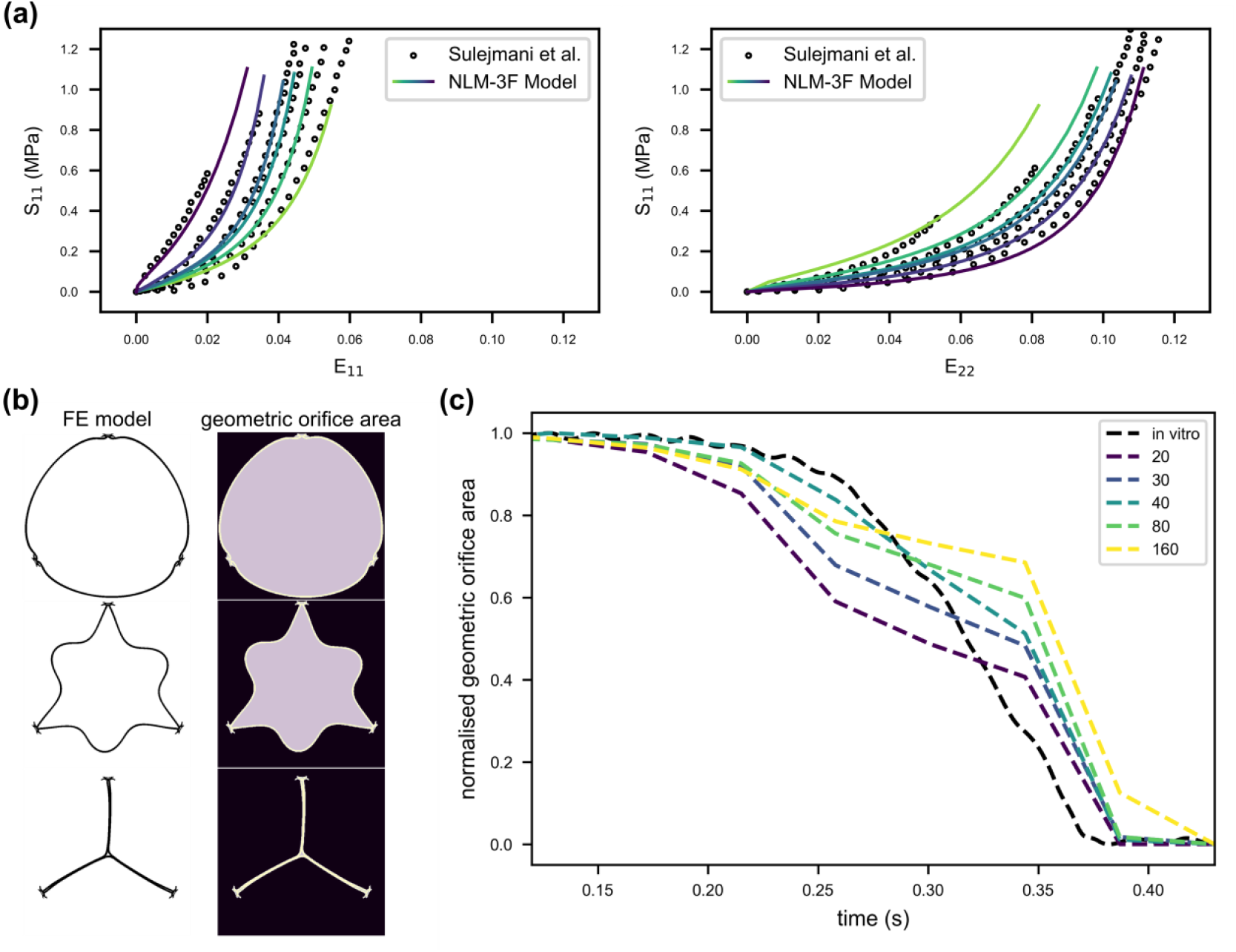
Material constitutive modeling of glutaraldehyde-fixed porcine pericardium. (a) Non-linear matrix, 3-fibre (NLM-3F) model fit to multi-protocol biaxial data obtained from Sulejmani et al. (2019). (b) Calculation of geometric orifice area (GOA) for sensitivity analysis. (c) Sensitivity study on the impact of material damping and device dynamics during the cardiac cycle.

Glutaraldehyde-fixed porcine pericardium was assumed to have a uniform density of 1,140 kg/m^3^ [11]. We conducted a sensitivity analysis on the impact of the mass proportional damping coefficient, α, on leaflet dynamics, by applying various α coefficients ranging from 1/s to 40/s to the leaflet [31]. As described in Section 2.2.2 below, we simulated the cardiac cycle through the application of hypertensive pressure boundary conditions. We calculated the geometric orifice area (GOA) throughout the cardiac cycle using a python code and compared results to those obtained from the validation experiments (Figure 2(b)). We found that the mass proportional damping coefficient did not notably impact the peak GOA of the leaflet during systole, but did however, influence on the kinematics of the leaflet during the period between peak systole and diastole (0.120 < t < 0.430 s) (Figure 2(c)). Based on this analysis, for all further models we applied a Rayleigh damping coefficient (α = 40 /s) for glutaraldehyde-fixed porcine pericardium to account for a variety of (tissue and blood) viscous effects [18, 26] and prevent un-physiological oscillations of the leaflet.

##### 2.1.1.2 Nitinol

To capture the superelastic, phase-transforming properties of nitinol, we used the constitutive law proposed by Auricchio and Taylor, which has been commonly used in previous studies of cardiovascular medical devices [11, 35]. Material constants were determined from tensile and compressive testing carried out at Boston Scientific, Galway, Ireland (Table 2). The material was assumed to have a density of 6,450 kg/m^3^. Following a sensitivity study, a damping coefficient (α = 35 /s) was used to dampen oscillations in the structure.

**Table 2:** Experimentally determined material parameters for nitinol.

| Parameter | Value | Units |
| --- | --- | --- |
| Austenite Youngs Modulus | 54.44 | GPa |
| Austenite Poisson Ratio | 0.3 |  |
| Martensite Youngs Modulus | 26.91 | GPa |
| Martensite Poisson Ratio | 0.3 |  |
| Transformation Strain | 0.047 |  |
| Start of Transformation Loading | 350 | MPa |
| End of Transformation Loading | 375 | MPa |
| Start of Transformation Unloading | 152 | MPa |
| End of Transformation Unloading | 120 | MPa |
| Start of Transformation during Compression Loading | 520 | MPa |
| Volumetric Transformation Strain | 0.047 |  |

### 2.2 Structural and hemodynamic simulation framework

#### 2.2.1 COU-1: Crimping and deployment

To simulate crimping and deployment, we aligned rigid plates concentrically around the THV. Two sets of 12 rigid plates were used for device crimping and deployment (upper and lower). These plates were positioned above and below the upper crown of the device (Figure 1(b)) respectively. Each rigid plate plane was discretized with discrete rigid elements (R3D4), with a global element size of 0.75 mm. Radial displacement boundary conditions were applied to each rigid plate to crimp to THV to an outer diameter of 10 mm (Figure 3). To prevent rotation and axial displacement of the device during crimping, a number of nodes on the stent at the lower crown, upper crown and stabilization arches (Figure 1(b)), uniformly arranged around the circumference, were held with zero azimuthal and axial displacement. A frictionless general contact, comprising “hard” contact with the penalty friction formulation was defined between each of the contacting surfaces involved in the crimping procedure, including the rigid contact plates, stent, leaflets, inner skirt and outer sealing skirt surfaces. The crimping process was simulated in a single timestep of 0.30 s.

**Figure 3:**
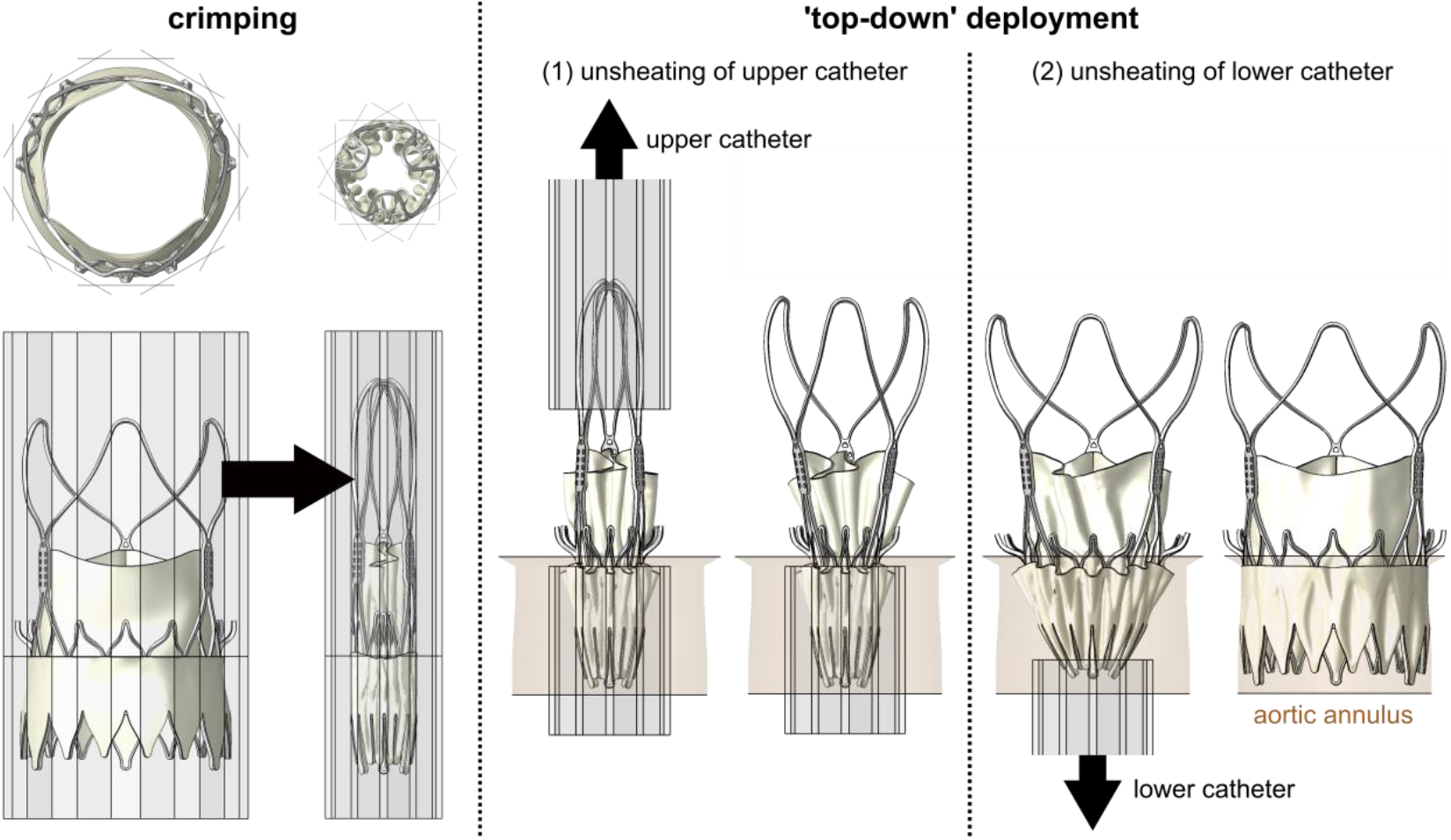
Finite element modeling of crimping and ‘top-down’ deployment of ACURATE Prime XL THV.

THV deployment was simulated using a two-step process, to replicate the unique ‘top-down’ deployment mechanism of the ACURATE Prime XL. This involved axial displacement of the 12 upper rigid plates in a single timestep (0.30 s), and axial displacement of the 12 lower plates in a single subsequent timestep (0.30 s), see Figure 3. During unsheathing of the lower plates, frictional “hard” contact with a friction coefficient of 0.20 was defined between the THV and rigid aortic annulus surfaces, which were meshed with discrete rigid elements (R3D4) with a global element size of 0.50 mm.

To validate across a range of device deployment configurations, the THV was deployed within a range of different annuli geometries. As shown in Table 3, the annulus models were characterized by the oversizing and frame ellipticity index, relative to the size of the ACURATE Prime XL (29 mm - recommended for annulus perimeter derived diameters ranging from 26.5 mm to 29 mm). Indices were calculated using the following equations from Dvir et al. [36] and Ruck et al. [37] respectively:

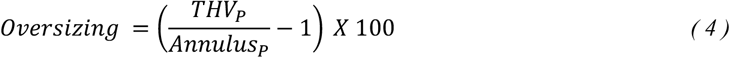

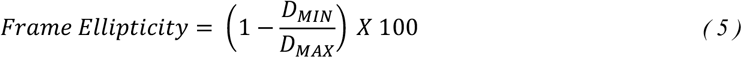

where *THV*_*P*_ and *Annulus*_*P*_ is the perimeter of the device (29 mm) and annulus respectively, and *D*_*MIN*_ and *D*_*MAX*_ are the minimum and maximum annulus diameters respectively. In this study, we conducted validation across a range of oversizing and ellipticity indices ranging from 0-10% and approximately 0-25% respectively, which aligns with indices measured across a wide range of patient cohorts in various clinical studies [37, 38]. Based on these calculations, deployment configurations for the THV were classified as the following: (1) ‘min-oversizing, circular’, (2) ‘max-oversizing, circular’ and (3) ‘min-oversizing, elliptical’.

**Table 3:** Relative expansion and ellipticity indices relative to ACURATE Prime XL.

|  | Min-oversizing,<br>circular | Max-oversizing,<br>circular | Min-oversizing,<br>elliptical |
| --- | --- | --- | --- |
| Perimeter-derived<br>diameter of annulus<br>($Annulus_P$) | 29 mm | 26.5 mm | 29 mm |
| Oversizing | 0 % | 9.4 % | 0 % |
| Frame Ellipticity | 0 % | 0 % | 24 % |

#### 2.2.2 COU-2: Cardiac cycle device dynamics

Once the device was deployed within a rigid aortic annulus model, we simulated the cardiac cycle by employing the general contact algorithm between the stent, leaflets, inner skirt and outer sealing skirt and aortic annulus surfaces, comprising “hard” contact with the penalty friction formulation and a friction coefficient of 0.20. The prescribed friction coefficient prevents stent sliding within the holding fixture, whilst also ensuring representative leaflet-to-leaflet interaction [39]. Hypertensive physiological pressure waveforms, extracted from a ViVitro pulse duplicator (described in Section 2.3.3), operating at a pressure waveform replicating a cardiac cycle of 70 BPM (0.86 s) with a peak trans-aortic pressure of 140 ± 3 mmHg, were applied to the aortic and ventricular surfaces of the leaflets. Following a cycle variance study, in which we simulate three hypertensive cardiac cycles and calculate the variance in stent deflection, we extracted results from the second cardiac cycle, given that it is representative of a periodic cardiac cycle. The complete FE simulation for each case, comprising crimping, deployment (COU-1) and cardiac cycles (COU-2), were solved using 128 CPU cores (2.6 GHz AMD Rome CPUs), with wall clock times ranging from 28 - 32 hours.

#### 2.2.3 COU-3: Cardiac cycle device hemodynamics

To analyze hemodynamics across the device during the cardiac cycle, we performed CFD simulations using OpenFOAM-v2312, where the device geometry at peak systole obtained from the FE simulations described above was integrated into an independent fluid flow simulation. The flow domain geometry was based on the geometry of the ViVitro pulse duplicator used for *in vitro* testing of the THV, described in Section 2.4.2 and Section 2.4.3, where the inlet and outlet was extended by approximately three times the length of the annulus diameter in order to avoid boundary effects on the flow field [14, 40]. The computational grid was discretized using native OpenFOAM mesh utilities ‘blockMesh’ and ‘snappyHexMesh’, whereby the meshing process occurs in two-distinct stages. Initially, a background mesh is created around the device and flow domain, which is then adjusted to conform to the boundary of the flow domain through cell-splitting, removal and surface snapping. Further mesh refinement was performed around the device to capture each component, whilst also ensuring a balance between computational cost and solution accuracy. Following a mesh sensitivity analysis examining the impact of global and local flow parameters, each flow domain consisted of 2.8 – 3.3 million hexahedral cells (Figure 4(b)).

**Figure 4:**
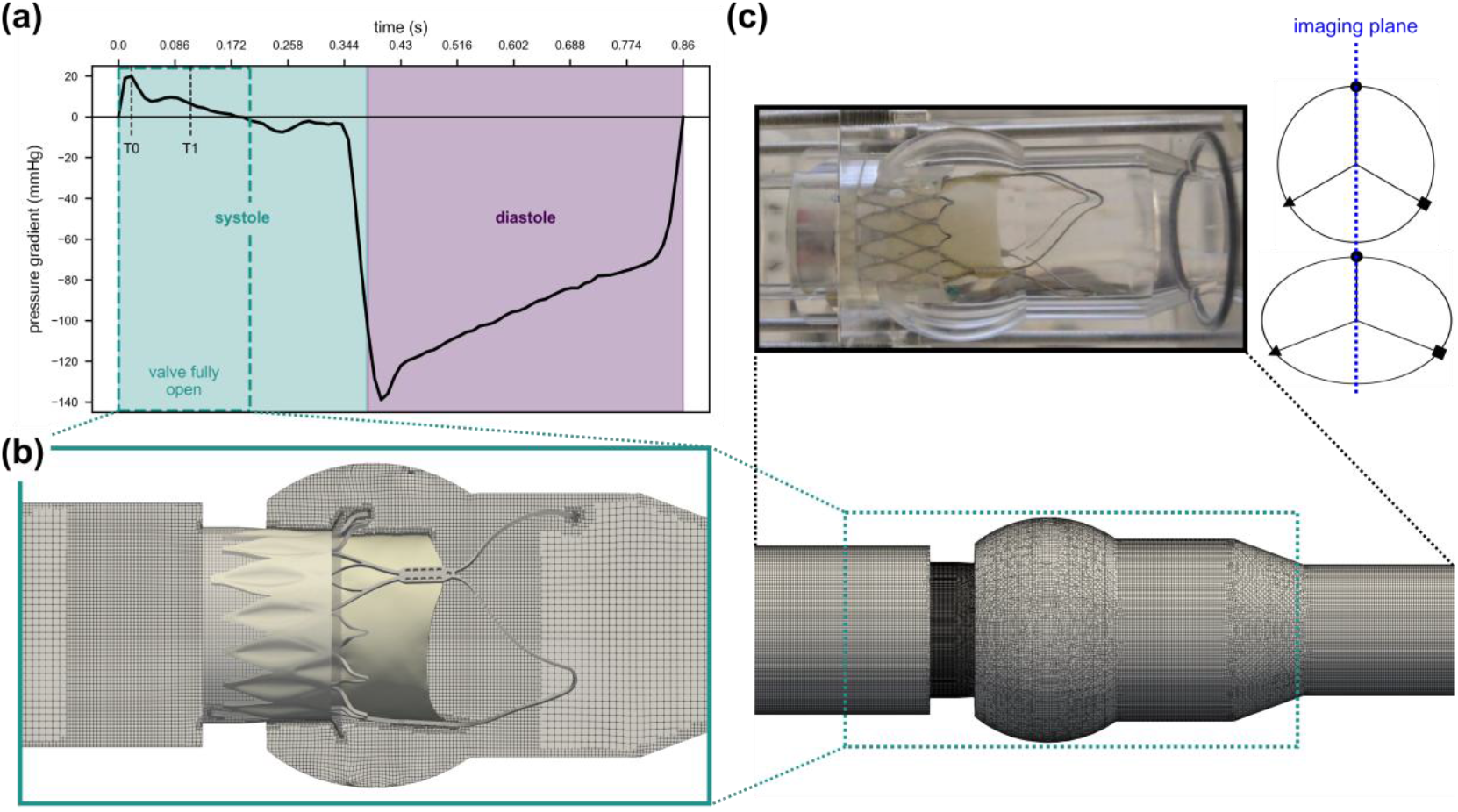
Modeling and validation of THV hemodynamics. (a) Pressure gradient applied at inlet showing early systole (T0) and peak systole (T1). (b) Mesh detail of systolic flow model. (c) Experimental PIV set-up showing imaging plane and corresponding in silico flow model.

**Figure 5:**
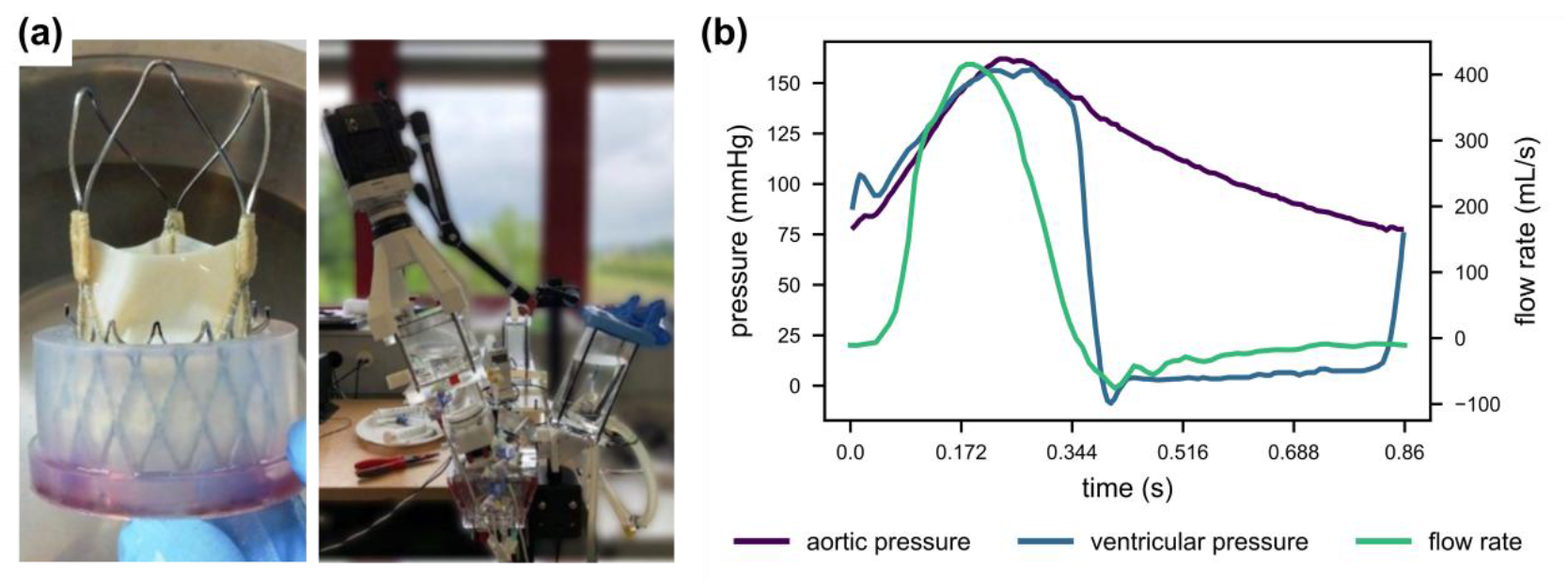
Structural high-speed image testing of THV. (a) ACURATE Prime XL and ViVitro pulse duplicator aortic root chamber with high-speed camera. (b) Moderate hypertensive cardiac cycle conditions.

Transient flow analyses were performed in OpenFOAM using the PISO-SIMPLE (PIMPLE) algorithm, where solutions were converged to a normalized residual of 1 x 10^-5^ at each time step. Temporal discretization was achieved using a second-order backward implicit scheme [41]. Convection terms were discretized using a second-order accurate linear upwind scheme, whilst gradients were computed using a second-order Gauss linear scheme. Fluid properties were representative of saline-glycerol solution, which was assumed to be Newtonian and incompressible with a density of 1120 kg/m^3^ and viscosity of 3.8 cP. For each deployment configuration, a systolic flow analysis was performed by the application of a time-dependent pressure gradient (Figure 4(a)), similar to previous approaches [42, 43]. All structures were treated as rigid, non-permeable, and a no-slip boundary condition was applied at the walls. The models were solved for 3 cardiac cycles, whereby a dynamic time-stepping scheme was chosen to ensure the Courant-Friedreich-Lewy (CFL) number was below 1.0 at all times.

### 2.3 Verification

As part of the verification studies, we performed as recommended [30]: (1) crimping and expansion simulations of the THV stent to a diameter of 10 mm (Figure 6(b)), to investigate the impact of (a) element formulation and (b) target time increment on local and global parameters relating to COU-1, and (2) simulations of cardiac cycle device dynamics to investigate the impact of element formulation on stent fatigue life predictions. Note that in a previous study [10], we conducted a mesh sensitivity analysis to determine spatial discretization and mesh quality error. Table 4 provides a summary of the COUs under study, with details of the verification and validation activities performed.

**Table 4:**
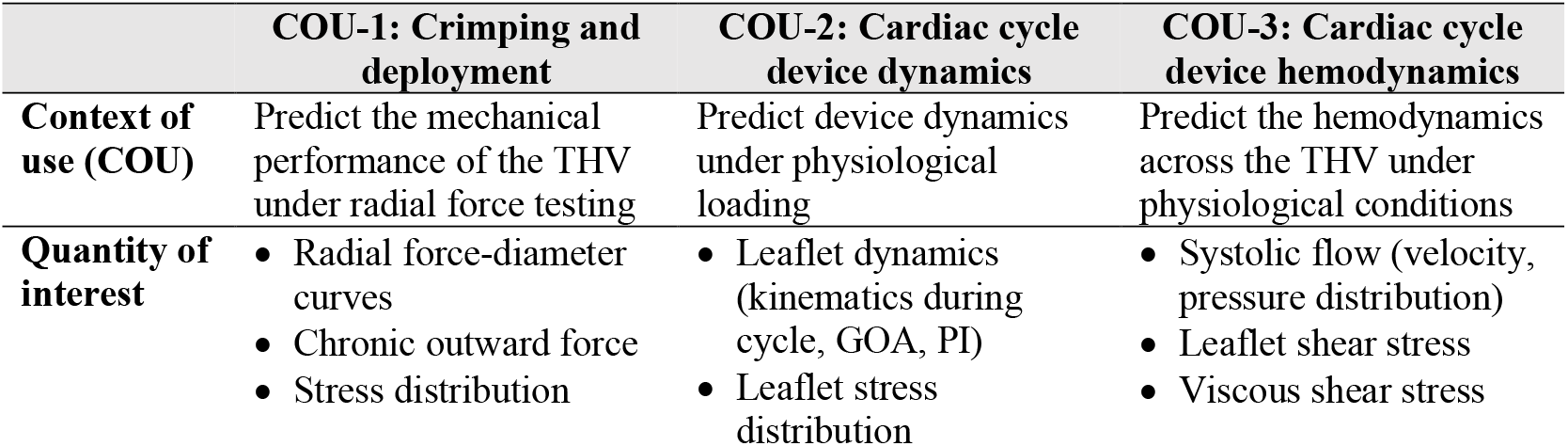
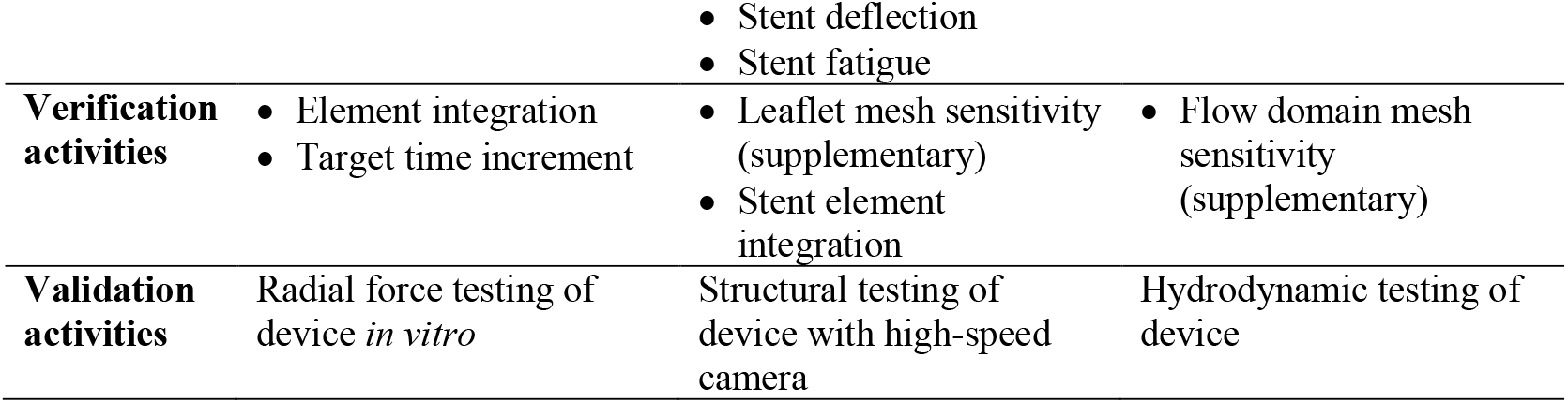
Context of use’s (COUs) of the THV model with relevant verification and validation activities.

|  | COU-1: Crimping and deployment | COU-2: Cardiac cycle device dynamics | COU-3: Cardiac cycle device hemodynamics |
| --- | --- | --- | --- |
| **Context of use (COU)** | Predict the mechanical performance of the THV under radial force testing | Predict device dynamics under physiological loading | Predict the hemodynamics across the THV under physiological conditions |
| **Quantity of interest** | • Radial force-diameter curves<br>• Chronic outward force<br>• Stress distribution | • Leaflet dynamics (kinematics during cycle, GOA, PI)<br>• Leaflet stress distribution | • Systolic flow (velocity, pressure distribution)<br>• Leaflet shear stress<br>• Viscous shear stress |
|  |  | • Stent deflection<br>• Stent fatigue |  |
| **Verification activities** | • Element integration<br>• Target time increment | • Leaflet mesh sensitivity (supplementary)<br>• Stent element integration | • Flow domain mesh sensitivity (supplementary) |
| **Validation activities** | Radial force testing of device *in vitro* | Structural testing of device with high-speed camera | Hydrodynamic testing of device |

**Table 5:** Results of the calculation verification on element formulation relating to context of use (COU)-1; THV crimping.

| Element formulation | Verification |  |  |  |  |
| --- | --- | --- | --- | --- | --- |
|  | Red-A | Full | Red-AH | Red-O | Red-C |
| CPU time | C | 3.65C | 0.86C | 0.88C | 0.92C |
| Radial resistive force at maximum crimp (N) | 61.59 | 72.38 | 84.59 | 61.89 | 67.50 |
| % Difference | / | +17.5% | +37.4% | +0.5% | +9.6% |
| Chronic outward force at 26.5 mm (N) | 18.74 | 22.24 | 26.53 | 18.66 | 19.70 |
| % Difference | / | +18.7% | +41.6% | -0.4% | +5.1% |
| Stress at maximum crimp (MPa) | 936.66 | 1612.98 | 982.52 | 932.67 | 890.95 |
| % Difference | / | +72.2% | +4.9% | -0.4% | -4.9% |

**Figure 6:**
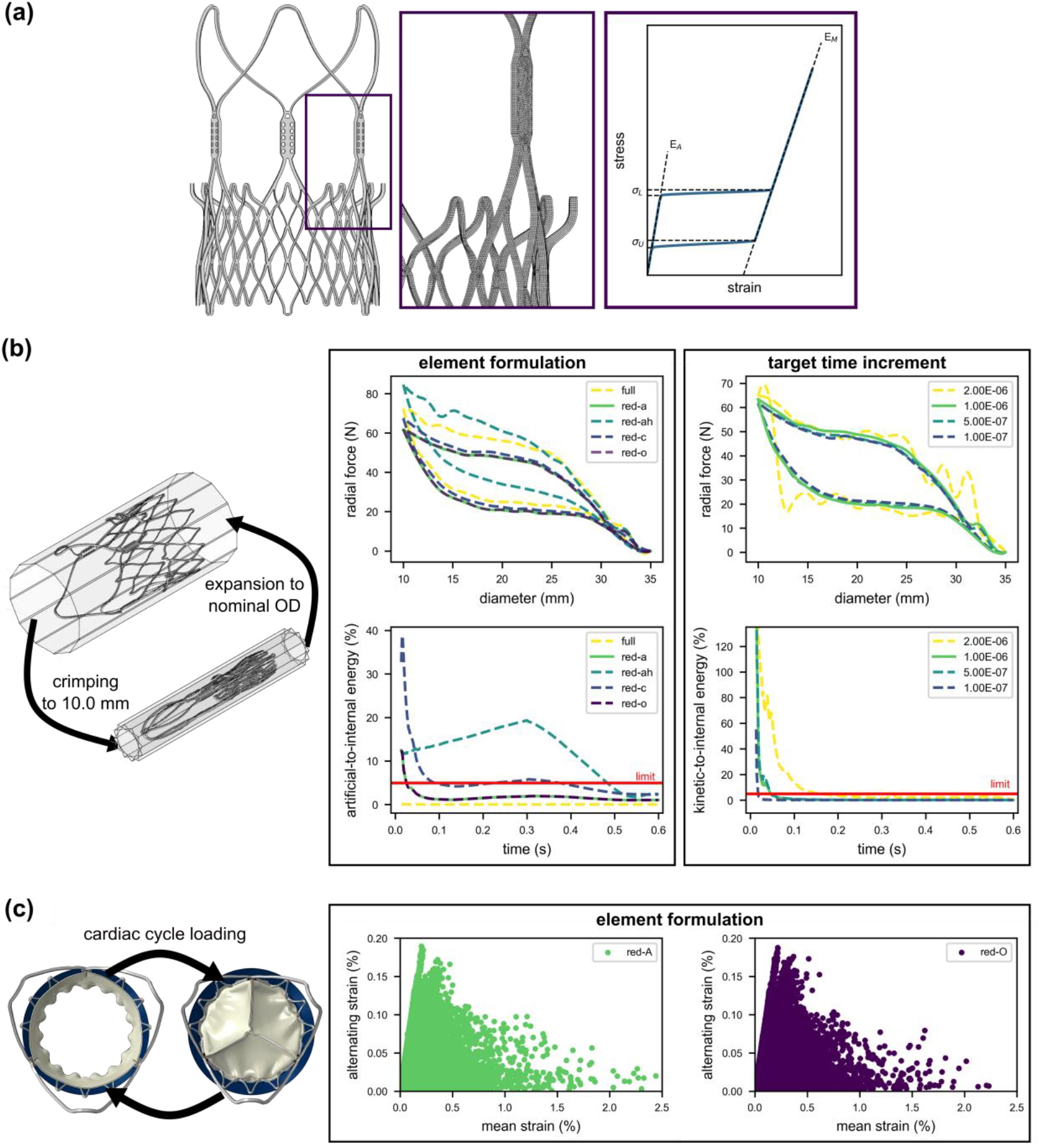
Calculation verification of THV model. (a) Numerical model of THV stent showing mesh detail and nitinol material model. (b) Verification of context of use (COU)-1: crimping and expansion simulation (left), showing the impact of full-integration (full), default reduced integration (red-a), reduced integration with enhanced hour glass control (red-ah), reduced integration with orthogonal kinematic split (red-oh) and reduced integration with centroid kinematic split (red-o), element formulations on the radial force and ratio of artificial-to-internal energy (middle), the impact of target time increment on the radial force and ratio of kinetic-to-internal energy (right). (c) Verification of COU-2: cardiac cycle device dynamics revealed the impact of element formulation on the alternating and mean strain.

In this study, we used three-dimensional linear solid elements to discretize the stent. To address the influence of using different mathematical element formulations on the impact of crimping and deployment, we examined the impact of using full and reduced-integration options. Within the reduced formulation, we also examined the impact of various kinematic split options (average (default), orthogonal and centroid) and enhanced hourglass control. During these simulations, we used a target time increment of 5 x 10^-7^. We also examined the impact on outputs relating to the context of use (COU) and analyzed the ratio of the artificial energy (includes energy stored in hourglass resistance and transverse shear) to internal energy, which should remain below 5%. Similarly, we examined the impact of the element formulation on cardiac cycle device dynamics and in particular, stent fatigue predictions, where we simulated the cardiac cycle as described in Section 2.2.2 above. From these simulations, we obtained the alternating strain (*ε*_*a*_) and mean strain (*ε*_*m*_) at each element in the THV stent, which is calculated as described in [32].

All FE simulations were performed using Abaqus/Explicit (v2023, Simulia, Dassault Systémes, Providence, RI, USA) due to the complex, non-linear contact conditions in the crimping, deployment and cardiac cycle simulation. In this study, a semi-automatic mass scaling strategy was applied throughout the step, every 1,000 increments, by selecting an element target time increment, with values of 2 x 10^-6^ s, 1 x 10^-6^ s, 5 x 10^-7^ s and 1 x 10^-7^ s for a total step time of 0.3 s. We examined the impact of target time increment on common outputs relating to THV stent crimping and the ratio of kinetic energy to internal energy, which should remain below 5% to satisfy quasi-static criteria.

### 2.4 Model Validation

#### 2.4.1 COU-1: Crimping and deployment

To validate the crimping and deployment simulations, and the nitinol material model, we conducted radial force testing of the ACURATE Prime XL inflow section (Figure 7(a)) at a controlled temperature of 37 °C. This involved crimping of a THV (n = 15) using a radial force tester to a nominal diameter of 23.5 mm; the mechanism was then reversed to allow for device re-expansion to a nominal diameter of 29 mm. The chronic outward force acting on the radial tester was measured at a nominal outer diameter of 26.5 and 29 mm. This test method was replicated *in silico* using the numerical model of the THV as described in Section 2.2.1 above and compared to the *in vitro* data. *In silico*, the radial force is calculated by obtaining the sum of the nodal radial reaction forces acting on each of the rigid plates, where the resistive radial force refers to the radial force during crimping, whilst the chronic outward force refers to radial force during stent expansion.

**Figure 7:**
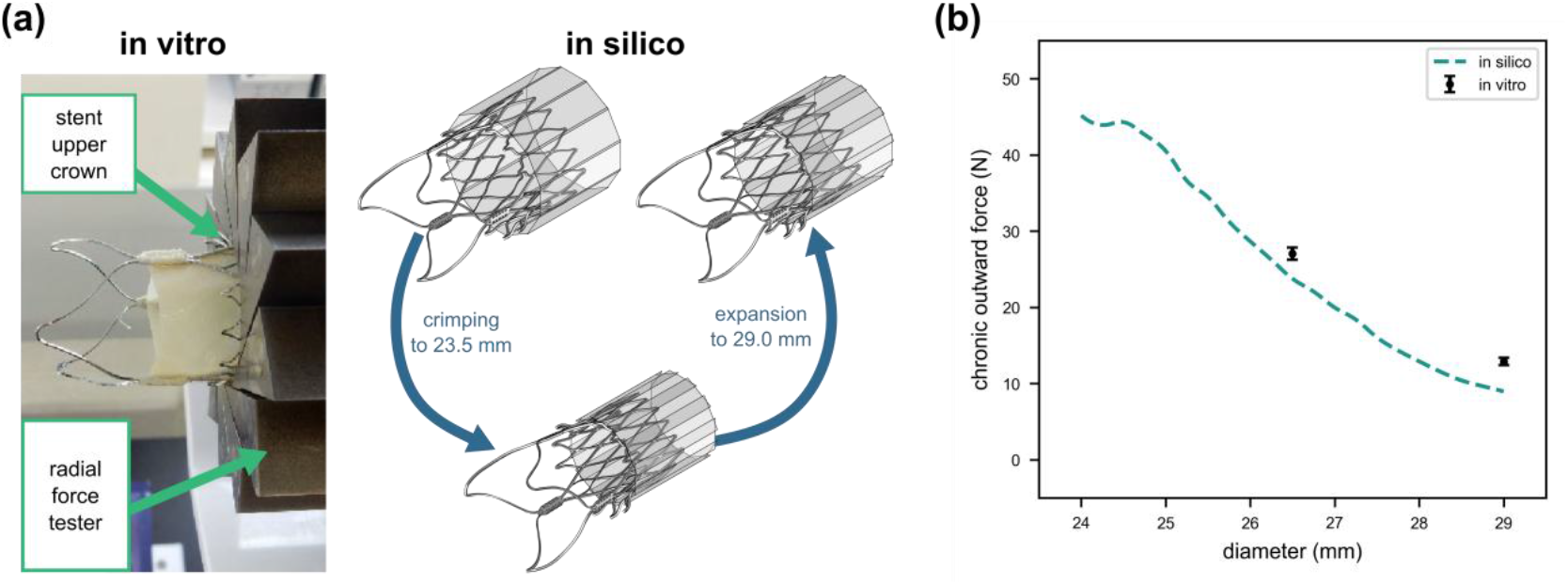
Validation of crimping and deployment (context of use 1) with radial force testing. (a) Radial force testing of ACURATE Prime XL inflow section. (b) Chronic outward force from radial force testing at 26.5 mm and 29 mm, with simulation results.

#### 2.4.2 COU-2: Cardiac cycle device dynamics

To validate FE models of the cardiac cycle dynamics, we conducted *in vitro* testing of the THV using a ViVitro pulse duplicator (ViVitro Labs Inc., Victoria, Canada). To capture THV dynamics used for validating the developed FE model, we mounted a high-speed camera above the aortic root chamber (Figure 5(a)). The device was then deployed within the aortic annuli models described in Section 2.2.1, and classified as; (1) ‘min-oversizing, circular’, (2) ‘max-oversizing, circular’ and (3) ‘min-oversizing, elliptical’. The deployed device was then installed within an aortic root chamber, which was operated at pressure waveforms replicating typical moderate hypertensive cardiac cycles of 70 BPM (0.86 s), with a peak trans-aortic pressure of 140 ± 3 mmHg (Figure 5(b)). The high-speed camera captured device dynamics, recording images at a frame rate of 200 fps and a resolution of 2560 × 1600. Using an in-house code, we calculated the geometric orifice area (GOA) over the cardiac cycle (n=1), which was used to validate leaflet motion. Further image processing was used to calculate the pinwheeling index (PI) at peak diastole (n = 1), an indicator of leaflet durability, using the following [44]:

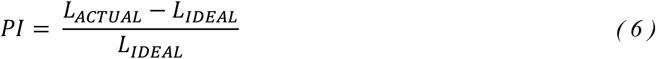

where *L*_*ACTUAL*_ represents the deflected free edge of the leaflet and *L*_*IDEAL*_ represents the ideal configuration of the leaflet free edge. We also calculate stent deflection, a measure of the radial deflection of each stent commissure (n = 12) between systole and diastole.

#### 2.4.3 COU-3: Cardiac cycle device hydrodynamics

To capture bulk flow patterns for validating the developed CFD model, we also performed hydrodynamic testing utilizing particle image velocimetry (PIV), an optical-based method for flow visualization. Similar to structural testing described above, the device was deployed within a ‘min-oversizing, circular’, ‘max-oversizing, circular’ and ‘min-oversizing, elliptical’ annulus, and the pulse duplicator operated at cardiac conditions with a normal cardiac output of 5 L/min, as shown in Figure 5(b). The PIV system consisted of a Nd:Yag pulsed laser (Litron Systems Ltd, Rugby, UK; Nano S 65-15 PIV, 65 mJ energy, 532 nm wavelength, 8 ns pulse duration), which was used to illuminate FluoSpheres™ polystyrene particles (ThermoFisher Scientific, MA, USA; Ø = 10 µm, emission at 560 nm) seeded in the fluid volume. Velocity maps in the immediate vicinity of the THV were acquired across the PIV imaging plane (Figure 4(c)), whereby image pairs were acquired every 0.054 s (16 phases per cardiac cycle) and repeated 40 times for each phase per cardiac cycle.

## 3 Results

### 3.1 Verification

#### 3.1.1 Element formulation

To investigate the impact of stent element formulation, we assumed reduced integration hexahedral elements (Red-A) as the reference model, as reduced integration is suggested for hexahedral elements applied to bending problems [45]. We found that the use of the centroid kinematic split (Red-C) resulted in the largest deviations of all outputs relating to COU-1; the resistive force at maximum crimp increased by 10%, the chronic outward force increased by 5% and the maximum stress at maximum crimp decreased by 5%, when compared to the reference element formulation. The ratio of energy dissipated as artificial-to-internal strain energy also surpassed maximum recommendations of 5% [45]. The use of orthogonal kinematic split (Red-O) resulted in similar COU-1 outputs, whilst satisfying both artificial-to-internal energy ratio requirements, and reducing computational cost by 12%, when compared to reference element formulation. The use of the enhanced hourglass control (Red-AH) option, resulted in large increases in all COU-1 outputs, whilst also increasing computational cost. For this element formulation, the ratio of artificial-to-internal strain energy exceeded 20%. The use of the full integration element (Full) resulted in an overly stiff material response, thereby causing a large increase in all global and local outputs relating to the resistive force at maximum crimp (+18%), the chronic outward force (+19%) and the stress at maximum crimp (+72%), whilst also increasing computational cost by 365%. Based on this analysis, we examined the impact of using the reference element formulation and orthogonal kinematic split on stent fatigue life predictions during the cardiac cycle (related to COU-2) but found no notable difference (Figure 6(c)).

#### 3.1.2 Target time increment

We assumed the reference target time increment to be 1 x 10^-7^. For all analyses, a peak of kinetic-to-internal energy ratio was found at the beginning of the simulation when the crimping plates initially contact the TAVI stent. After this initial peak, the ratio of kinetic energy to internal energy was below 5% across all analyses, thereby satisfying quasi-static criteria [45]. As shown in Table 6, the target time increment had a minimal impact on the force and stress at maximum crimp. The chronic outward force at 26.5 mm decreased by approximately 15.7%, 6.2% and 4.3% for a target time increment of 2E-6, 1E-6 and 5E-07 respectively, when compared to the reference target time increment.

**Table 6:** Results of the calculation verification on target time increment relating to context of use (COU)-1; THV crimping.

| Target time increment | Verification |  |  |  |
| --- | --- | --- | --- | --- |
| | $2 \times 10^{-6}$ s | $1 \times 10^{-6}$ s | $5 \times 10^{-7}$ s | $1 \times 10^{-7}$ s |
| CPU Time | 0.05C | 0.10C | 0.21C | C |
| Radial resistive force at maximum crimp (N) | 64.63 | 63.38 | 61.59 | 62.05 |
| % Difference | +4.2% | +2.2% | -0.8% | / |
| Chronic outward force at 26.5 mm (N) | 16.50 | 18.37 | 18.74 | 19.58 |
| % Difference | -15.7% | -6.2% | -4.3% | / |
| Stress at maximum crimp (MPa) | 970.36 | 937.02 | 936.66 | 924.53 |
| % Difference | +5.0% | +1.4% | +1.3% | / |

### 3.2 Model validation

#### 3.2.1 COU-1: Crimping and deployment

Radial force testing results for the chronic outward force during expansion of the THV were 27.5 ± 0.8 and 12.9 ± 0.5 N at 26.5 and 29 mm respectively. The simulation of this experimental set-up was well aligned with experimental data, predicting a chronic outward force of 24.0 and 9.0 N at 26.5 and 29 mm respectively (Figure 7(b)).

#### 3.2.2 COU-2: Cardiac cycle device dynamics

The FE modeling framework was validated across (1) ‘min-oversizing, circular’, (2) ‘max-oversizing, circular’ and (3) ‘min-oversizing, elliptical’ deployment configurations, with corresponding supplementary videos (Supplementary Materials Video S1-S3) showing device dynamics during the cardiac cycle compared with structural image testing. As evident from Figure 8, device dynamics throughout the cardiac cycle were similar to those captured through high-speed imaging of the *in vitro* experiment. The peak GOA *in silico* was within a target error requirement of 10% for each deployment configuration. It is important to note, that, similar to previous studies [23], our model slightly overpredicted GOA. This may be an artefact of using the FE method, which approximates forward fluid flow by imposing a normal pressure distribution across the surface of the leaflets. In the ‘min-oversizing, circular’ deployment configuration, our FE model predicted a pinwheeling index (PI) of 2.68%, which was similar to that observed *in vitro* (2.62%). Our model also captured pinwheeling trends, also observed during *in vitro* testing, where pinwheeling increased in the ‘max-oversizing, circular’ annulus, compared to the ‘min-oversizing, circular’ and ‘min-oversizing, elliptical’ annuli.

**Figure 8:**
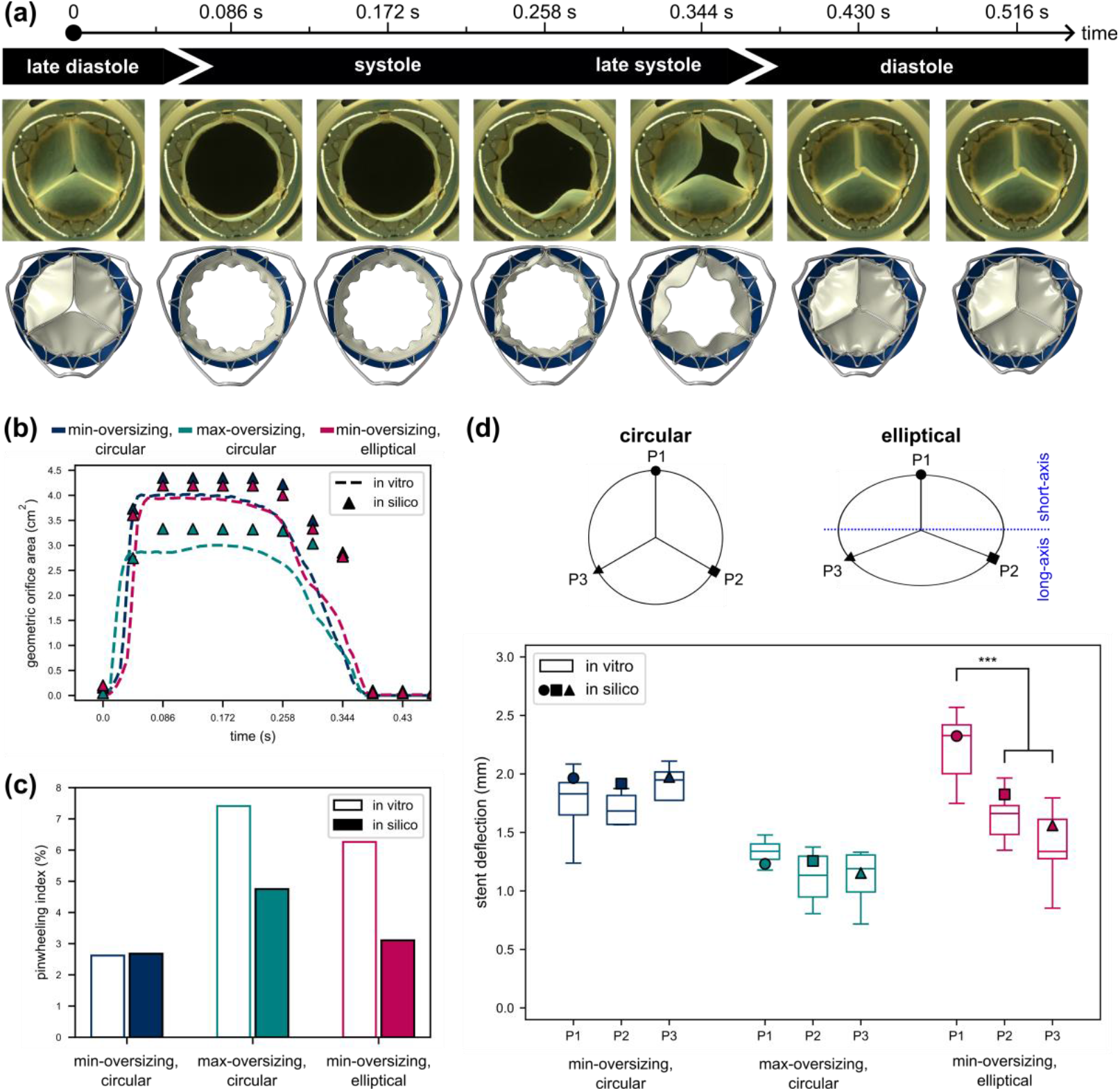
Validation of finite element model with structural imaging testing. (a) Device dynamics during the cardiac cycle as observed in in vitro testing (top) and FE model (bottom). (b) Geometric orifice area (GOA) during systole. (c) Pinwheeling index (PI) at peak diastole. (d) Stent deflection during cardiac cycle.

Finally, as observed in Figure 8(d), our model shows excellent alignment with stent deflection across each of the deployment configurations, whereby all predictions lay within one standard deviation of *in vitro* testing. This measurement is obtained by calculating the peak radial distance between the commissure posts at systole and diastole. The THV stent exhibited notable radial deflections (> 2 mm in some cases), due to the pulsatile loading of the tissue leaflets. As also observed in *in vitro* testing, our model predicts that stent deflection is approximately equal across commissure posts P1, P2 and P3, in circular deployment configurations. Of note, deployment of the device in the ‘max-oversizing, circular’ annulus reduced average stent deflections (1.21 mm) by over 35%, when compared to ‘min-oversizing, circular’ annulus (1.95 mm). In the ‘min-oversizing, elliptical’ annulus, maximum stent deflection occurs at commissure post P1, which is positioned along the short-axis of ellipticity. Commissure posts P2 and P3 (located along the long-axis of ellipticity) each exhibited deflections that were equal to approximately 80% of the deflection of commissure post P1 in the ‘min-oversizing, elliptical’ deployment configuration.

#### 3.2.3 COU-3: Cardiac cycle device hemodynamics

The CFD models were validated against PIV testing in the ‘min-oversizing, circular’, ‘max-oversizing, circular’ and ‘min-oversizing, elliptical’ annuli. Figure 9(a) shows the flow velocity contours across the device at early systole (t = 0.02 s) and peak systole (t = 0.12 s) in the ‘min-oversizing, circular’, ‘max-oversizing, circular’ and ‘min-oversizing, elliptical’ annulus for the experimental PIV and computational CFD model. At early systole, in each deployment configuration, a jet emerges from the valve orifice, which also generates low velocity vortices (between approximately 0.25 and 0.50 m/s) on either side. PIV shows that the maximum velocity within this jet just upstream of the THV is equal to 1.093 m/s, 1.014 m/s and 1.084 m/s for the ‘min-oversizing, circular’, ‘max-oversizing, circular’ and ‘min-oversizing, elliptical’ annulus respectively. As shown in Figure 9(a), our CFD model overpredicts the length of this jet stream at early systole, likely due to the fact that the valve remains fully open throughout the systolic flow simulation. At peak systole, the centralized jet continues to emerge from the valve orifice and the annular vortex generated during early systole moves gradually downstream of the THV. The maximum velocity is 1.273 m/s, 1.194 m/s and 1.230 m/s for the ‘min-oversizing, circular’, ‘max-oversizing, circular’ and ‘min-oversizing, elliptical’ annulus at peak systole respectively. As observed in the ‘min-oversizing, elliptical’ configuration, in both the PIV and CFD model, the high velocity flow jet is slightly asymmetric, with higher velocities tending towards the right of the imaging plane. This results in the formation of a recirculating zone to the left of the imaging plane, just upstream of the device, which again is clearly visible in both the PIV and CFD model. Of note, the CFD models show excellent alignment with maximum velocity predictions in both the ‘min-oversizing, circular’ and ‘min-oversizing, elliptical’ annuli at both early and peak systole, with relative errors of less than 10%. Our CFD model overpredicts the maximum velocity in the jet at both early and peak systole in the ‘max-oversizing, circular’ annulus, with complete results shown in Table 7. Throughout both early and peak systole, flow within the aortic sinus remains low (< 0.1 m/s) across *in vitro* testing and CFD models.

**Table 7:** Validation of CFD model with PIV hydrodynamic testing.

|  |  | Min-oversizing,<br>circular | Max-oversizing,<br>circular | Min-oversizing,<br>elliptical |
| --- | --- | --- | --- | --- |
| Maximum velocity @<br>early systole | *In vitro* (m/s) | 1.093 | 1.014 | 1.084 |
|  | *In silico* (m/s) | 1.185 | 1.229 | 1.158 |
|  | % Error | +8.42% | +21.20% | +6.83% |
| Maximum velocity @<br>peak systole | *In vitro* (m/s) | 1.273 | 1.194 | 1.230 |
|  | *In silico* (m/s) | 1.326 | 1.381 | 1.302 |
|  | % Error | +4.16% | +15.66% | +5.85% |

**Figure 9:**
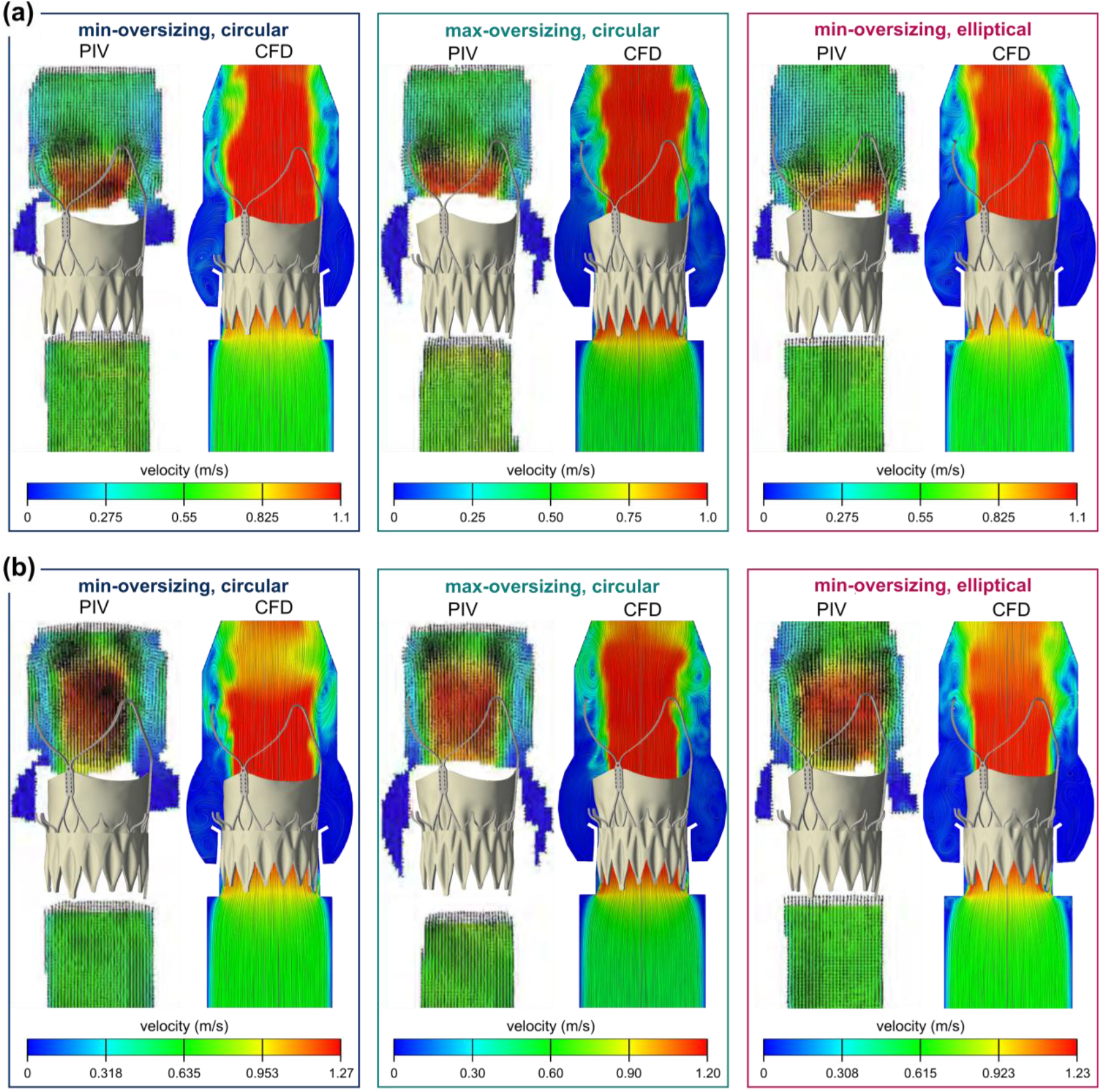
CFD model validation with particle image velocimetry (PIV) hydrodynamic testing at (a) early systole and (b) peak systole.

## 4 Discussion

In this study, we developed an advanced computational structural and hemodynamic framework for assessing the performance of a self-expanding THV. This framework utilized discrete computational solid mechanics and CFD modeling approaches, where the THV device model incorporated a novel advanced material model of glutaraldehyde-fixed porcine pericardium, a deformable nitinol stent with both inner and outer pericardial skirts. Based on ASME VV 40-2018, we conducted a range of verification and validation activities relating to (COU-1) crimping and deployment, (COU-2) cardiac cycle device dynamics and (COU-3) cardiac cycle device hemodynamics. We showed how these COUs are sensitive to numerical parameters including mesh density, element formulation and target time increment, which should be considered while developing computational models of THVs. Numerical model validation was conducted against radial force, structural high-speed imaging and hydrodynamic PIV testing. Overall, our models showed excellent alignment with *in vitro* testing across multiple cases of THV deployment, showing similar radial force, GOA, pinwheeling, stent deflection and flow velocity profiles. Thus, we established a workflow, encompassing all stages of the device crimping and deployment lifecycle, for verification and validation of TAVI simulations and demonstrated the credibility of our novel framework to predict the *in vitro* performance of a self-expanding THV.

There are some limitations worth considering. Firstly, we did not account for FSI between the device and adjacent fluid flow, but applied a hydrodynamic pressure using FE modeling and then exported the deformed device geometry for CFD modeling, thereby assuming rigidity. Nonetheless, we demonstrated the credibility of our modeling framework to accurately predict device performance across the cardiac cycle. Ongoing work aims to address this limitation by developing a fully-coupled FSI model of the self-expanding THV, whereby further verification and validation as per ASME VV-40 will be required. Secondly, as part of our CFD models, we assumed that fluid flow was laminar. Although intraventricular flow past THVs may undergo periodic transformations to laminar-transitional in certain phases of the cardiac cycle, previous studies have reported minor differences in near-wall flow when comparing the use of laminar and turbulent flow models [40]. Finally, as per ASME VV-40, uncertainty quantification may be used for specific model inputs [27, 46], to account for variances that may arise from manufacturing dimensional tolerances, material properties and experimental boundary conditions. Although outside the scope of the current work, future work may account for such uncertainties by utilizing statistical methods, such as Monte Carlo simulations, to examine how model uncertainties impact output predictions.

A strength of the current study is the implementation of (1) a novel non-linear matrix, three-fiber (NLM-3F) strain-stiffening model, to capture the anisotropic and highly non-linear behavior of the glutaraldehyde-fixed porcine pericardium, and (2) a deformable stent. This is based on our previous approach, which aimed to more accurately reproduce the complex material behavior of glutaraldehyde-fixed porcine pericardium over the full functional range, when compared to single-fiber models of soft tissue such as the HGO model [10]. This allows for precise control of the fiber-matrix stiffness ratio at small and large deformations, which has a notable impact on THV deformation. Previous studies utilizing similar strain-stiffening matrix and fiber models have provided more physical representations of soft tissue deformations [21, 47, 48]. Furthermore, previous computational studies of THVs have typically assumed that the THV stent frame is rigid, even though our experimental and computational model predicted that self-expanding THVs exhibit notable deformations due to pulsatile cardiac loading [10]. Here, we integrated an advanced constitutive leaflet model, a deformable superelastic nitinol stent and incorporated both the inner and outer pericardium skirts. This advanced model showed excellent alignment with *in vitro* testing.

The results of the calculation verification study revealed that both element formulation and target time increment impacted all global (radial resistive force at maximum crimp, chronic outward force at device working range) and local outputs (maximum stress at maximum crimp) relating to THV device crimping and deployment. Interestingly, the use of the enhanced hourglass control option, resulted in large increases in the radial resistive force at maximum crimp and the chronic outward force, whilst also increasing computational cost. Although such element formulation options are widely used in the literature [11, 49], in our case, the use of enhanced hourglass control resulted in substantial artificial energy, whereby the ratio of artificial-to-internal strain energy was well above the recommended 5% maximum limit. Based on these findings, the default integration or orthogonal kinematic-split options are recommended for three-dimensional linear solid elements of nitinol THV stents as they provide the most appropriate device response relating to the COUs investigated in this study. These findings are similar to those recently reported by Bernini et al. for crimping, deployment and fatigue prediction of a femoral artery stent [30]. In previous studies, the stability of explicit solutions schemes, such as that used in this study, are assumed if the solution satisfies general quasi-static conditions (ratio of kinetic-to-internal energy < 5%) [11-13]. Despite this, substantial deviations in the crimping and expansion behavior of femoral artery stents with varying target time increments have recently been reported in the literature [30]. In this study, we report similar findings for a self-expanding THV. Based on this analysis, it is worth noting that we employed a target time increment of 1 x10^-6^ s for the study to provide the optimal balance between computational cost and solution accuracy. We recommend future studies to conduct similar sensitivity analyses, irrespective of whether quasi-static requirements are met.

A number of previous studies have validated outputs related to commonly investigated COUs relating to THVs, including (1) crimping and deployment [13, 50], (2) cardiac cycle dynamics [19, 23, 43] and (3) cardiac cycle hemodynamics [43]. However, to the best of our knowledge, this is the first study to validate these COUs simultaneously, whilst also doing this across multiple THV deployment configurations. State-of-the-art bench testing was used to validate model outputs including radial force, structural and hydrodynamic testing, thereby providing a novel framework for validating THV performance. Our computational prediction of chronic outward force shows good alignment with radial force testing, which thereby validates COU-1 in relation to this study. Secondly, experimental data obtained for device dynamics during the cardiac cycle through a high-speed camera, validates GOA, pinwheeling and stent deflection, across a number of different deployment configurations. Specifically, stent deflection predictions were all within one standard deviation of *in vitro* testing. Finally, bulk flow predictions from CFD models were validated using hydrodynamic PIV testing, which was conducted in a ViVitro pulse duplicator, again predicting flow velocity within 5% of *in vitro* testing.

In part two of this comprehensive computational study, we utilize the validated device model to investigate the impact of device expansion and ellipticity on biomechanical device markers of degeneration [Reference to Part II]. Moreover, while the current study focuses on the verification and validation of the THV models, it is important to note the models developed in this study may be used for informing next-generation device design and pre-operative planning of patient-specific TAVI procedures.

## 5 Conclusion

Given the increasing use of *in silico* models and digital twins in healthcare, establishing the credibility of such models is of utmost importance. ASME-VV-40 documentation provides a risk-informed credibility assessment for numerical models of medical devices, which may be achieved through systematic verification and validation approach. In this study, we developed a novel *in silico* framework to assess the structural and hemodynamic *in vitro* performance of a supra-annular, self-expanding THV, which encompassed all stages of device lifecycle, from crimping to implantation. To the best of our knowledge, this is the first computational study to evaluate the robustness of a computational framework to address all major COUs relating to THVs, including (COU-1) crimping and deployment, (COU-2) cardiac cycle dynamics and (COU-3) cardiac cycle hemodynamics, utilizing both FE analysis and CFD. Through the integration of *in silico* modeling and comprehensive *in vitro* testing, which included radial force measurement, high-speed camera imaging and hydrodynamic PIV testing, we provide a novel framework for assessing the credibility of a FE and CFD-based modeling approach as outlined by ASME VV-40.

## Data Availability

All data produced in the present study are available upon reasonable request to the authors

## Acknowledgements

The authors would like to acknowledge the Irish Centre for High-End Computing (ICHEC) for provision of computational facilities and support.

