## Supplementary Materials for "In Silico Modeling of Transcatheter Heart Valve Oversizing and Ellipticity, Part I: Establishing Credibility of an Advanced Model"

#### 1 Leaflet mesh sensitivity study

To determine the optimal number of elements through thickness of the leaflet, we conduct a mesh sensitivity study, where models of the tri-leaflet component were created with 1, 2, 3 and 5 elements through the thickness. Each model was subject to moderate hypertensive physiological loads as described in Section 2.2.2 to simulate the cardiac cycle. The von Mises stress at peak diastole (t = 0.430 s) was defined as the target value for the sensitivity, where similar to previous approaches by Li et al., as shown in Figure S1, we define six locations located across the leaflet, which will be used to compare the von Mises stress across the different meshes. Based on this analysis, as presented in Figure S1, three elements through thickness of the leaflet were determined to provide an optimal balance between accuracy and computational cost, thus were used for the remainder of the study.

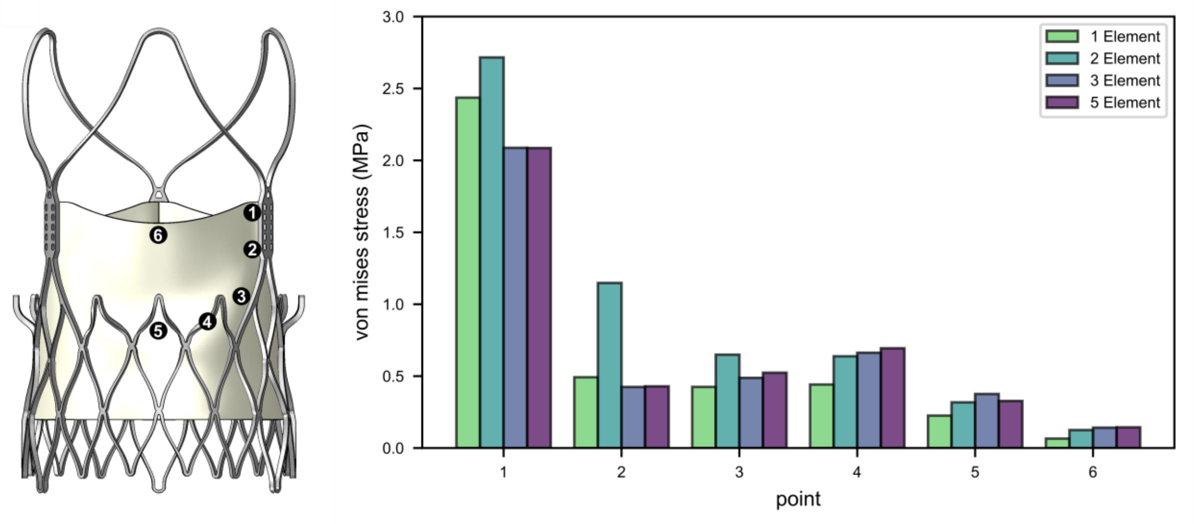


**Figure S1: Verification on the number of elements through thickness of the leaflet**

### 2 CFD mesh sensitivity study

#### 2.1 Bulk flow – *blockmesh*

A mesh independence study was performed on the CFD systolic flow domain using OpenFOAM, to evaluate the impact of discretization error on bulk flow predictions. This involved varying the average element size using OpenFOAM meshing utility *blockMesh*, whilst surface snapping (*snappyHexMesh*) remained constant, as shown in Table S1.

*Table S1: blockmesh refinement study for bulk flow*

| Mesh | blockMesh average cell size | snappyHexMesh level of refinement | Number of cells | Relative CPU time |
|---|---|---|---|---|
| **Coarse** | 2.00 mm | 1 | 208,444 | 0.1 C |
| **Intermediate** | 1.00 mm | 1 | 951,160 | C |
| **Fine** | 0.50 mm | 1 | 4,580,100 | 9.1 C |

In order to estimate the discretization error, we obtain the velocity at the inflow, valvular and outflow level, as shown in Figure S2. Next, we obtain the grid convergence index (GCI), which represents a 95% confidence interval for the estimated error using velocity values and equations 1-4:

$$p = \frac{ln\left(\frac{f_3 - f_2}{f_2 - f_1}\right)}{\ln(r)} \tag{1}$$

$$E_1^{fine} = \frac{f_2 - f_1}{1 - r^p} \tag{2}$$

$$GCI_1^{fine} = 1.25\left|E_1^{fine}\right| \tag{3}$$

$$f_{true} = f_1 \pm GCI_1^{fine} \tag{4}$$

where $f_{true}$ represents the 95% confidence interval for the true mesh independent value. The confidence interval for the peak velocity at the inflow, valve and outflow level is represented by the red-dashed lines in Figure S2(b), where results indicate that the intermediate mesh (average element size = 1 mm) is suitable for capturing bulk flow across the TAVI device.

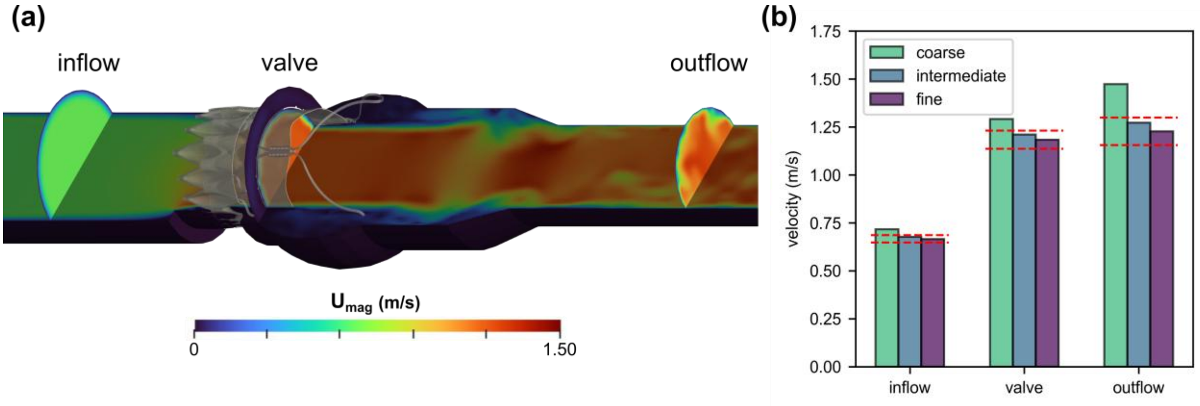


**Figure S2: blockMesh refinement study,** (a) Velocity contours at peak systole showing inflow, valve and outflow plane used to calculate doppler velocity. (b) Velocity at inflow, valve and outflow showing grid convergence index (GCI) confidence interval.

##### 2.1.1 Local flow – *snappyhexmesh*

A further mesh independence study was performed on the CFD systolic flow to evaluate the impact of discretization error on local flow predictions. This involved varying the level of surface snapping using OpenFOAM meshing utility *snappyHexMesh*, whilst the initial background mesh, created using *blockMesh*, remained constant with an average element size of 1 mm.

| Mesh | blockmesh average cell size | snappyhexmesh level of refinement | Number of cells | Relative CPU time |
|---|---|---|---|---|
| **Coarse** | 1.00 mm | 2 | 2,430,978 | X C |
| **Intermediate** | 1.00 mm | 3 | 2,805,157 | C |
| **Fine** | 1.00 mm | 4 | 8,627,468 | X C |

To estimate the discretization error on local flow parameters, we plot the time-averaged wall shear stress (TAWSS) on the aortic and ventricular surface of the leaflets throughout systole as shown in Figure S3. Following this discretization study, we use the intermediate mesh settings for further CFD models with a three levels of surface edge refinement to achieve the optimal balance between computational cost and solution accuracy.

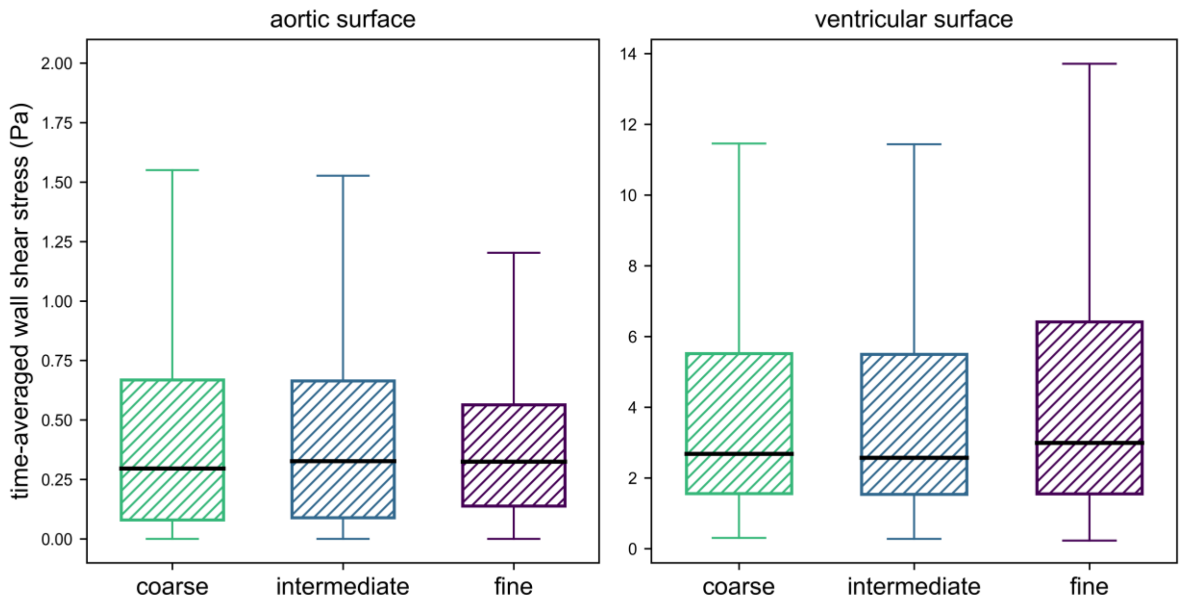


**Figure S3: snappyHexMesh refinement study** showing TAWSS on the aortic (left) and ventricular (right) surface of the leaflets throughout systole
